# Neuropathophysiological changes associated with mirtazapine treatment response in major depressive disorder: insights into bottom-up dopaminergic pathways and prefrontal control networks

**DOI:** 10.1101/2025.11.24.25340752

**Authors:** Younghwa Lee, Egle Simulionyte, Sandi Hebib, André Manook, Rosanne Picotin, Somayeh Mohammadi Joyandeh, Thomas C. Baghai, Jens Schwarzbach, Rainer Rupprecht, Oliver Gruber

## Abstract

Due to the heterogeneity in symptoms and underlying biological mechanisms of major depressive disorder (MDD), as well as the lack of biologically validated approaches for subtyping, effective treatment often requires a prolonged trial-and-error process. This heterogeneity underscores the need for objective biomarkers that can predict individual treatment responses and guide antidepressant selection. Although mirtazapine is widely used and its efficacy is well established in clinical settings, neuroscientific studies on its treatment response remain limited relative to other antidepressants. To address this gap, the present study investigated pathophysiological changes in MDD patients that were associated with subsequent treatment response to mirtazapine, using functional magnetic resonance imaging with a conditioned reward task. Sixty-seven individuals diagnosed with MDD were enrolled and treated with either mirtazapine or one of two comparator antidepressants: agomelatine or a selective serotonin reuptake inhibitor. Participants were classified as responders or non-responders after six weeks of treatment.

Mirtazapine responders demonstrated increased pre-treatment activation in core components of the mesolimbic dopaminergic pathway, particularly the ventral tegmental area and ventral striatum, as well as in prefrontal and functionally connected regions implicated in reward evaluation, reward-related behavioral inhibition, and reward-based decision-making. Importantly, many of these response-related activations were specific to the mirtazapine group.

These findings suggest that the observed activation patterns may reflect alterations in the mesolimbic dopaminergic pathway and prefrontal regulatory networks during reward processing, which may characterize the pathophysiological brain features of MDD patients who respond to mirtazapine treatment. Moreover, these activation patterns may serve as promising biomarkers for identifying an MDD subtype more likely to benefit from mirtazapine.

## 1. Introduction

Major depressive disorder (MDD) is a complex and increasingly prevalent psychiatric illness worldwide. Symptom expression in MDD is highly heterogeneous, exhibiting substantial interindividual variability (Fried and Nesse, 2015a; Nemesure et al., 2024). Moreover, individual depression symptoms may have different underlying biological correlates (Fried and Nesse, 2015b). However, due to the current diagnostic system’s reliance on symptom-based criteria alone and the absence of standardized guidelines for prescribing antidepressants based on biological or neurological evidence, predicting treatment outcomes for MDD remains a significant challenge. Consequently, approximately 60% of patients undergo a prolonged trial-and-error process before finding an effective treatment (Fava, 2003). This has led to an increasing interest in identifying biomarkers that can predict responses to antidepressant medications. Among these efforts, neuroimaging, particularly functional magnetic resonance imaging (fMRI), has emerged as a promising non-invasive tool to capture neuropathophysiological features associated with MDD subtypes and their treatment responses.

Mirtazapine, a widely used antidepressant, has a distinct pharmacological mechanism involving antagonism of adrenergic α_2_ receptors and blockade of serotonergic 5-HT_2_, 5-HT_3_, and histamine H_1_ receptors—unlike classical monoamine reuptake inhibitors, which primarily increase extracellular serotonin and/or norepinephrine levels by inhibiting their reuptake (Davis, 2020). This mechanism contributes to the alleviation of MDD symptoms by enhancing levels of serotonin, norepinephrine, and dopamine (either directly or indirectly) through α_2_ receptor antagonism and 5-HT_2_/5-HT_3_ blockade (Davis, 2020; Devoto et al., 2004). By blocking 5-HT_2_ and 5-HT_3_ receptors, mirtazapine may reduce selective serotonin reuptake inhibitor (SSRI)-related side effects such as anxiety and insomnia (Davis, 2020; Nutt, 1999), while its H_1_ receptor antagonism provides additional sedative benefits for patients with MDD (Davis, 2020). Despite its distinct pharmacological profile and well-established clinical benefits, only a limited number of neuroimaging studies have examined the neural correlates of treatment response to mirtazapine. Existing research has primarily focused on emotional processing regions (Frodl et al., 2011; Komulainen et al., 2017; Lisiecka et al., 2011; Rawlings et al., 2010), whereas other neural systems critically involved in MDD have received comparatively little attention.

One such system is the brain reward network, which also plays a critical role in the pathophysiology of MDD. Numerous studies have consistently shown an association between MDD and abnormal activation in brain regions involved in reward processing (Jiang et al., 2023; Ng et al., 2019; Oh et al., 2023).

These abnormalities manifest in various forms across patients, and the resulting functional patterns are thought to contribute to the heterogeneity observed in MDD, including the emergence of distinct subtypes with different pathophysiological profiles (Borsini et al., 2020; Foell et al., 2021; Foti et al., 2014; Goya-Maldonado et al., 2015; Jullig et al., 2024). In response, recent studies have employed reward-related fMRI tasks to identify neural markers that may characterize MDD subtypes and predict differential treatment responses (Greenberg et al., 2020; Nguyen et al., 2022; Nguyen et al., 2019; Tura and Goya-Maldonado, 2023). Despite this progress, brain regions involved in the reward system have been largely overlooked in studies examining treatment response to mirtazapine. To date, no studies have specifically explored the relationship between mirtazapine response and functional brain features related to reward processing in patients with MDD.

To address this gap, the present study aimed to investigate neuropathophysiological changes in patients with MDD that may be associated with treatment response to mirtazapine, with a specific focus on the extended reward system. We examined pre-treatment differences in neural activation between mirtazapine responders and non-responders using fMRI and a conditioned reward task, which measured responses to reward-related stimuli typically associated with bottom-up reward processing. To evaluate the specificity of these activation patterns, we also included comparison groups treated with other antidepressants, including agomelatine and SSRIs.

## 2. Experimental procedures

### 2.1 Study design

As part of the OptiMD-SP5 study’s naturalistic, bi-center study design, MDD inpatients undergoing antidepressant monotherapy were recruited between February 2016 and July 2021 at the Department of General Psychiatry, Heidelberg University, and the Department of Psychiatry and Psychotherapy, University of Regensburg. Inclusion criteria were MDD diagnosis based on the ICD-10 and DSM-V (first episode or recurrent), age 18–65 years, and written informed consent. Exclusion criteria were incapacity to provide informed consent, contraindications to MRI scanning, presence of an organic mental disorder (e.g., intoxication or CNS neurological disease), neurodegenerative disorders, relevant physical illnesses, pregnancy or breastfeeding, current or past substance dependence (i.e., clinical diagnosis of dependence), cannabis use in the past 2 weeks, and drug use (other than cannabis) in the past month.

Before initiating a new antidepressant monotherapy, patients underwent fMRI scanning while completing a conditioned reward task, the Desire-Reason-Dilemma (DRD) paradigm. A conditioning phase and a practice session first took place outside of the scanner. A compensation of 30 euros was given to subjects, who could further win a 20 euros bonus through the DRD task. Measurements with the Montgomery-Åsberg Depression Rating Scale (MADRS), the Hamilton Rating Scale for Depression (HDRS), and the Clinical Global Impression Scale (CGI) were conducted weekly for 6 weeks, and the antidepressant monotherapy was adjusted if necessary. Study approval was obtained from the responsible ethics committees.

### 2.2 Participants

A total of 121 patients with MDD were enrolled. Patients were treated with one of six antidepressant monotherapies: mirtazapine, agomelatine, SSRIs (either sertraline or escitalopram), SNRI, bupropion, or tricyclic antidepressants. Sixteen patients were in treatment groups with currently insufficient numbers to be included in analyses (SNRI = 11, bupropion = 3, tricyclic antidepressants = 2), and 38 patients were excluded during quality control (comedication and/or comorbidity = 21, study discontinuation and/or unclear treatment response status = 14, poor fMRI scan quality = 3). The final sample included 67 patients, comprising 23 patients in the mirtazapine group, 18 in the agomelatine group, and 26 in the SSRI group. Patients with at least four weeks of clinical measurements were included in the analyses. Response classification was conducted based on a comprehensive clinical evaluation, in which MADRS score improvement served as the primary criterion. Patients were initially classified into three preliminary categories according to mean MADRS score improvement during weeks 4–6 from the initial visit: <25% as non-responders, 25–50% as partial responders, and ≥ 50% as responders. Subsequently, HDRS scores, along with individual clinical profiles over time (checking for consistency) and information about the subsequent clinical course (after week 6), were taken into consideration for a more in-depth clinical judgment of treatment response. This process led to a final classification into treatment responders vs. non-responders. Patients whose treatment response data did not allow for an unambiguous judgement of treatment response were excluded from further analyses. All statistical analyses for demographic and clinical characteristics were performed using SPSS 29.0.2.0 (IBM, Chicago, Illinois, USA).

### 2.3 Experimental paradigm

The DRD paradigm was employed to investigate bottom-up responses to conditioned reward stimuli (Figure 1). Prior to the fMRI experiment, participants underwent a training session outside the scanner consisting of two distinct phases. In the first phase, participants completed an operant conditioning task in which eight colored squares were presented in random order (20 repetitions each). They freely responded by accepting (left button) or rejecting (right button) each stimulus using their right hand. Through trial-and-error feedback, two colors (red and green) were associated with reward (+10 points), four with a neutral outcome (0 points), and two with punishment (-10 points). Punishment-associated colors were included only in this first phase to prevent a response bias toward the left (accept) button in the second phase. This phase served to establish stimulus-reward associations.

**Figure 1.**
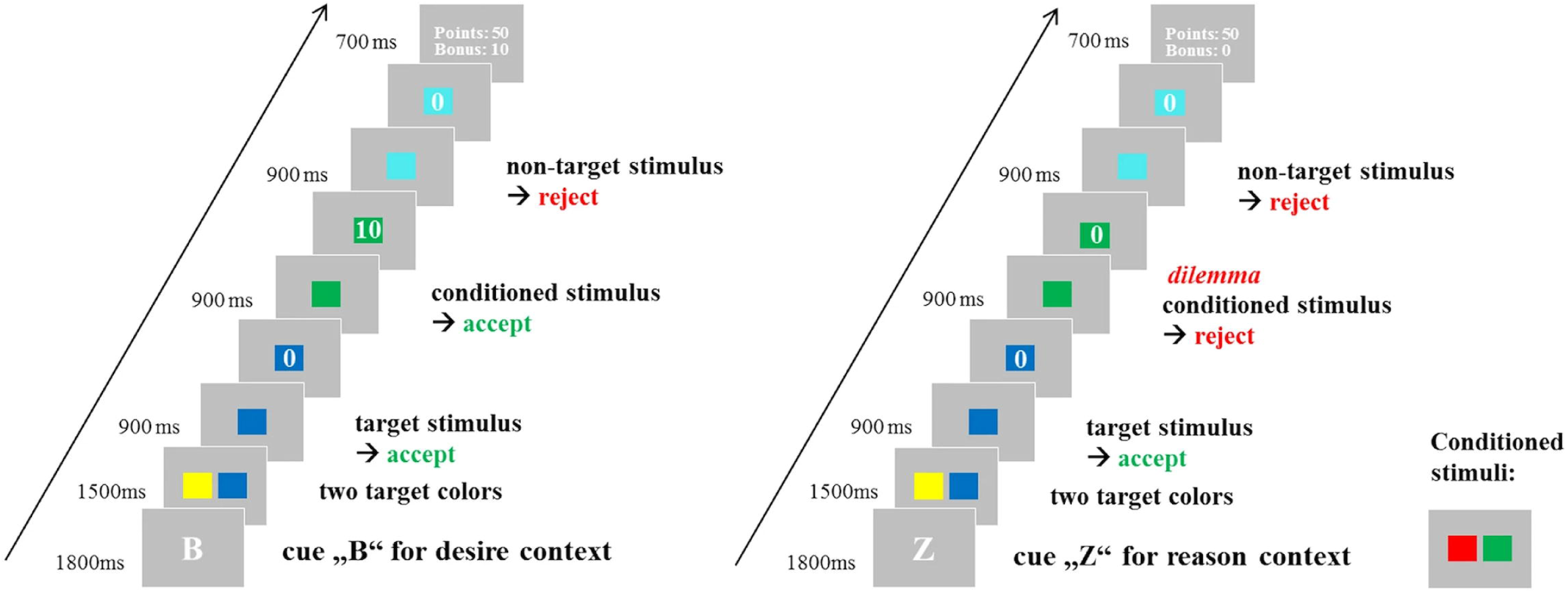
Desire-Reason-Dilemma paradigm. At the beginning of each sequence, target stimuli to be accepted were shown. In the desire context (left), participants were also prompted to accept previously conditioned reward stimuli to receive an additional 10-point bonus per stimulus. In the reason context (right), participants were instructed to reject those stimuli in order to obtain the superordinate 50 points at the end of each block.

In the second phase, participants were familiarized with the DRD paradigm, a sequential forced-choice task. Two types of blocks were used: the ‘desire context’ (DC) and the ‘reason context’ (RC). Each block consisted of four to eight trials, with each trial involving the presentation of a single-colored stimulus. Before each block, two target colors were shown out of four colors. Subsequently, participants were required to accept these colors throughout the block in order to achieve a superordinate goal of 50 points. In the DC blocks, previously conditioned reward stimuli could also be accepted to obtain additional bonuses of 10 points. In the RC blocks, this was not allowed, and erroneous acceptance of the conditioned stimuli led to the loss of the 50-point goal. More details were provided in our previous studies (Diekhof and Gruber, 2010; Diekhof et al., 2012).

On the day after the training session, participants underwent the actual experiment, performing 40 task blocks of the DRD paradigm during two fMRI runs. The points obtained in the scanner were converted into actual monetary rewards. For the present study, only the DC blocks—designed to capture bottom-up responses to conditioned reward stimuli—were included in the analysis.

### 2.4 fMRI

Bicentric MR scanning of patients was carried out at the Neuroradiology Department of Heidelberg University Hospital, and at the Clinical Neuroscience Campus of the University of Regensburg using Siemens 3T whole-body scanners. For the completion of the DRD task, the scanner was also equipped with a right-hand response keyboard and a mirror on the head coil for visual stimulation. A T2*-weighted echo-planar imaging sequence was acquired while patients completed the DRD task. Each of the overall two scanning sessions contained 185 volumes covering the whole brain, with TE = 30 ms and TR = 1900 ms for a total scan duration of around 6 min (flip angle = 70°, voxel size = isotropic 3 mm, matrix = 64 x 64, field of view = 192 x 192 mm, number of slices = 31 (axial), distance factor = 20%, acquisition sequence and direction = ascending from posterior to anterior). The first volumes of each session were discarded before preprocessing.

Raw fMRI data were converted from DICOM to NIfTI. The converted images were preprocessed and analyzed with Statistical Parametric Mapping 12 software (SPM, https://www.fil.ion.ucl.ac.uk/spm/software/spm12/). Preprocessing consisted of realignment and unwarping, correction for slice-time acquisition differences using the first slice as a reference, normalization into the Montreal Neurological Institute (MNI) stereotactic space, and spatial smoothing with a 6 mm full-width at half-maximum (FWHM) isotropic Gaussian kernel filter.

For the first-level analysis, a general linear model (GLM) was fitted to the data using standard procedures as described in Diekhof et al. (2012). Vectors representing the temporal onsets of different experimental and stimulus conditions were convolved with the canonical hemodynamic response function to produce the predicted hemodynamic response to each experimental condition. Linear t-contrasts were defined by contrasting activation effects elicited by the conditioned stimuli in the DC relative to the implicit baseline to examine reward-related activation patterns.

For the second-level analysis, a full factorial design was created using treatment arms (mirtazapine, agomelatine, and SSRI) and response groups (responders and non-responders) as factors. Contrasts were established to examine whole-brain activity differences between responders and non-responders for each treatment arm and to compare treatment arms against each other using interaction contrasts. The initial statistical search criterion for group statistics was *p* < .05, uncorrected.

Furthermore, since the DC of the fMRI task used in our analysis was designed to engage bottom-up reward processing, we paid particular attention to the activation in the bilateral ventral striatum (VS) and ventral tegmental area (VTA), regions that have consistently been identified as core components of the reward system in previous research. These regions (VS: ±12, 12, −3; VTA: ±9, −21, −12) were defined a priori based on coordinates reported in a previous study (Diekhof and Gruber, 2010), and a 3-mm radius sphere was applied to each for small volume correction to account for multiple comparisons. Statistical significance was assessed using a family-wise error rate (FWE)–corrected threshold of *p* < .05 within these volumes. This approach was chosen to optimize the detection of activation in the brain regions closely relevant to our research task, while also mitigating the overly conservative nature of whole-brain correction in fMRI data.

## 3. Results

### 3.1 Sample characteristics

A total of 67 participants were analyzed in this study, with a mean age of 30.8 years (SD = 11.8); 43.3% of the participants were female. According to the naturalistic study design, patients were assigned based on their clinical profile to one of three treatment groups: mirtazapine (n = 23), agomelatine (n = 18), or SSRIs (n = 26; sertraline = 7, escitalopram = 19). Except for gender, there were no significant differences in demographic or clinical characteristics among the treatment groups (Table 1). The proportion of female participants in the SSRI group (26.9%) was significantly lower than in the mirtazapine (43.5%) and agomelatine (66.7%) groups (*p* = .03). Across the total sample, the mean duration since the first depressive episode was 4.9 years (SD = 7.1), and the mean number of depressive episodes was 2.2 (SD = 3.8). At baseline, the mean MADRS score was 25.5 (SD = 5.3), which decreased to 15.6 (SD = 7.1) during weeks 4–6, corresponding to a mean reduction rate of 38.3% (SD = 7.1). Overall, 74.6% of participants (n = 50) were classified as responders, including 18 in the mirtazapine group, 13 in the agomelatine group, and 19 in the SSRI group. Comparative analyses among treatment groups for these demographic and clinical characteristics were conducted using only cases without missing data. Detailed demographic and clinical characteristics, as well as analysis results, are presented in Table 1.

**Table 1.** Demographic and clinical characteristics by each treatment group.

|  | All | Treatment arms |  |  | p-value |
| --- | --- | --- | --- | --- | --- |
|  |  | Mirtazapine | Agomelatine | SSRI |  |
| n | 67 | 23 | 18 | 26 |  |
| Age, years, mean (SD) | 30.8 (11.8) | 32.5 (12.4) | 34.3 (13.7) | 26.9 (8.57) | .08 |
| Females, n (%) | 29 (43.3) | 10 (43.5) | 12 (66.7) | 7 (26.9) | .03* |
| Years since first episode, mean (SD) / n | 4.9 (7.1)/60 | 4.0 (7.3)/22 | 4.9 (5.1)/16 | 5.8 (8.2)/22 | .73 |
| Number of episodes, mean (SD) / n | 2.2 (3.8)/59 | 1.6 (2.8)/20 | 2.9 (3.9)/16 | 2.2 (4.5)/23 | .57 |
| MADRS at baseline, mean (SD) | 25.5 (5.3) | 24.7 (4.5) | 25.1 (6.2) | 26.7 (5.2) | .38 |
| Mean MADRS (W4-W6), mean (SD) | 15.6 (7.1) | 16.2 (5.6) | 15.2 (7.6) | 15.4 (8.0) | .90 |
| MADRS reduction rate, %, mean (SD) | 38.3 (25.5) | 33.3 (22.9) | 39.2 (24.6) | 42.1 (28.3) | .48 |
| Maximum dose of antidepressant (mg), mean (SD) | - | 37.6 (8.0) | 47.2 (8.1) | Sert (n=7)<br>139.3 (47.6) | - |
|  |  |  |  | Esc (n=19) |  |
|  |  |  |  | 15.6 (3.7) |  |

Note. Values are presented as mean (SD) or number (%), unless otherwise indicated. p-values were calculated using one-way ANOVA for continuous variables and chi-square test for categorical variables. $p < .05$ was considered statistically significant. Asterisks (\*) indicate statistical significance at $p < .05$ . Mean MADRS score (week 4-6) was calculated as the average of available MADRS scores obtained between Week 4 and Week 6 for each participant. MADRS reduction rate was calculated as $[(\text{baseline MADRS} - \text{mean MADRS score (week 4-6)}) / \text{baseline MADRS}] \times 100$ for each participant. Maximum dose of antidepressant is presented in milligrams (mg) with separate values reported for sertraline and escitalopram within the SSRI group. Sample sizes (n) may vary across variables due to missing data. Abbreviations: MADRS, Montgomery-Åsberg Depression Rating Scale; SD, standard deviation; SSRI, selective serotonin reuptake inhibitor; Sert, sertraline; Esc, escitalopram

### 3.2 Activity differences in mirtazapine responders vs non-responders

Differences in neural activation between mirtazapine responders and non-responders were observed in the VTA and VS, which are core a priori regions involved in bottom-up reward processing, as well as in additional brain areas, including prefrontal, parietal, temporal, and insular cortices and the anterior thalamus. To determine whether these differences reflected task-relevant (i.e., reward-related) activity rather than non-task-related background signals, the identified regions were assessed for overlap with prototypical task-related activation as defined by a reference map in the DRD paradigm. This reference map was derived in SPM by performing a one-sample t-test on brain activation elicited by conditioned reward stimuli across a large sample of 224 healthy controls from the Genomic Imaging Göttingen (GIG) cohort, who had completed the same paradigm in a prior study (Trost et al., 2016; Wolf et al., 2016). Activation differences between mirtazapine responders and non-responders in regions aligning with prototypical reward-related activation defined by the reference map, representing the main findings of the present study, are listed in Table 2 and visualized in Figure 2. Further differences that did not overlap with the DRD activation reference map are briefly described in Supplementary Table 1.

**Figure 2.**
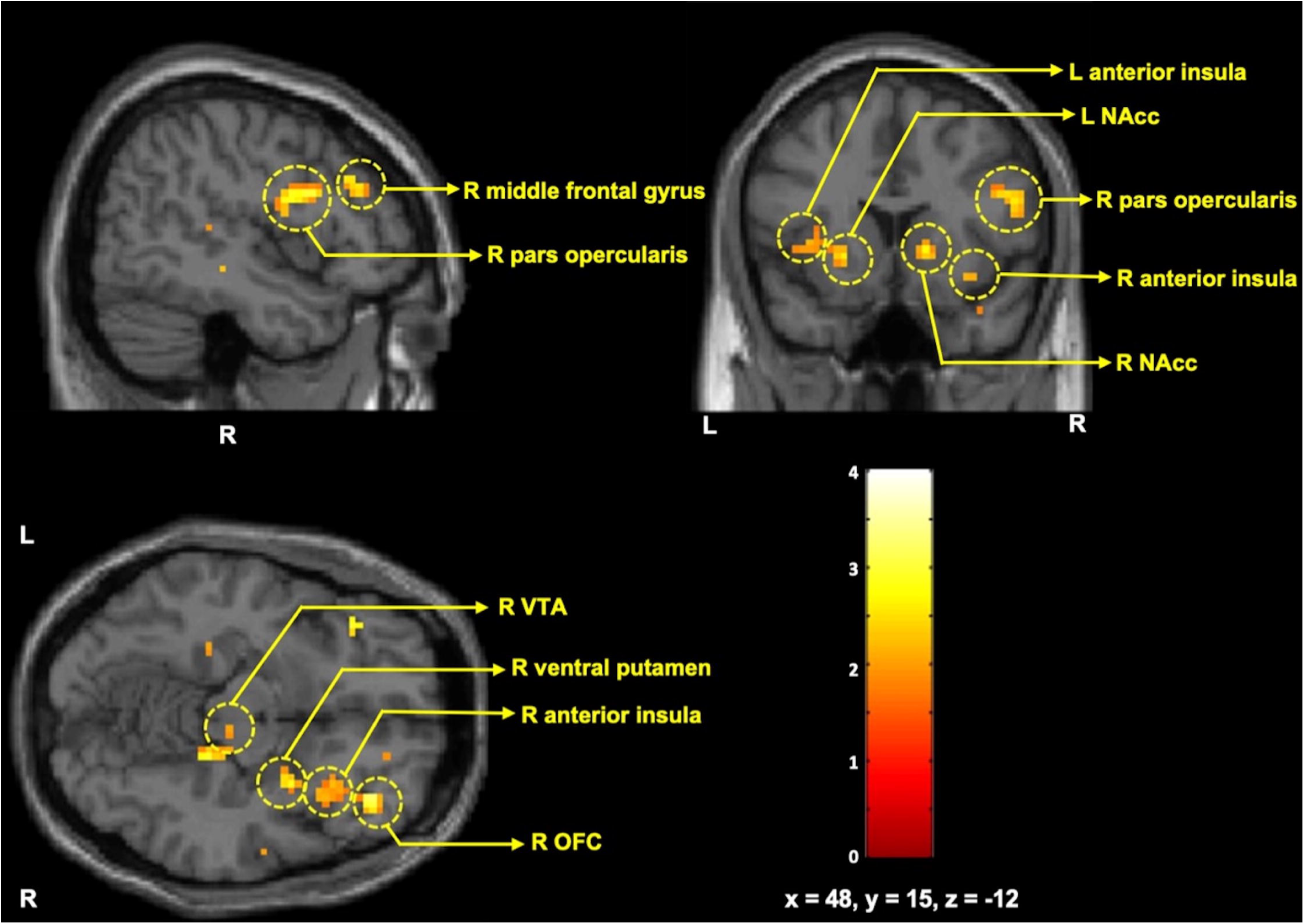
Statistical parametric map (T-map) showing relatively increased brain activity in response to conditioned reward stimuli in mirtazapine responders compared to non-responders (*p* < .05, uncorrected). Coordinates are reported in the Montreal Neurological Institute (MNI) stereotactic space. Abbreviations: L, left; R, right; VTA, ventral tegmental area; NAcc, nucleus accumbens; OFC, orbitofrontal cortex.

**Table 2.** Increased pre-treatment activity in responders to mirtazapine relative to non-responders.

| Region |  | MNI coordinate (T-value) |  |  |
| --- | --- | --- | --- | --- |
|  |  | Mir RS > NRS | Interaction contrast |  |
|  |  |  | Mir > Ago | Mir > SSRI |
| (A) Increased activity specifically associated with subsequent treatment response in Mir<br>(Not observed either in Ago or SSRI) |  |  |  |  |
| Cortical |  |  |  |  |
| Prefrontal | R IFG, pars opercularis | 48 3 24 (3.25) | 57 6 18 (2.38) | 51 9 24 (3.61) |
|  | R central OFC | 42 39 -12 (2.80) | 36 39 -12 (3.01) | 36 39 -9 (3.40) |
|  | R MFG, middle third | 51 33 24 (2.39) | 48 30 30 (2.14) | 51 33 24 (2.67) |
| Temporal | L fusiform gyrus | -36 -48 -15<br>(2.58) | -36 -51 -18<br>(2.67) | -36 -48 -15<br>(2.58) |
| Subcortical |  |  |  |  |
| VS | R ventral putamen | 30 3 -12 (3.32) | 30 3 -12 (2.99) | 30 3 -12 (1.98) |
|  | L anterior ventral putamen | -24 15 -3 (2.96) | -21 15 -3 (2.02) | -21 15 -3 (2.53) |
| VTA | R VTA | 12 -24 -9 (2.57)<sup>†</sup> | 6 -24 -12 (2.09) | 15 -24 -12<br>(2.61) |
| (B) Increased activity associated with response in Mir, but merely in a partially specific manner<br>(Not observed in SSRI, but comparable or lower in Ago) |  |  |  |  |
| Cortical |  |  |  |  |
| Prefrontal | R frontomedian cortex | 6 12 27 (2.90) | n.s. | 6 12 27 (3.12) |
|  | L posterior SFS | -24 -9 57 (2.41) | n.s. | -24 -9 57 (2.58) |
|  | R avPFC | 30 45 15 (2.18) | 33 48 15 (2.03) | 30 48 15 (2.79) |
| Parietal | L intraparietal cortex | -21 -45 48<br>(2.87) | n.s. | -24 -42 48<br>(2.63) |
| Temporal | L TOC | -36 -72 3 (3.03) | -36 -75 3 (1.84) | -36 -69 0 (2.53) |
| Insular | L anterior insula | -39 12 -3 (3.03) | -36 9 0 (1.73) | -36 6 3 (3.80) |
|  | R anterior insula | 33 18 -12 (2.85) | 33 24 -9 (1.76) | 33 15 -12 (2.72) |
|  | L posterior insula | -45 -3 6 (2.75) | n.s. | -42 -3 9 (3.15) |
| Subcortical |  |  |  |  |
| VS | R NAcc | 15 15 0 (3.14)* | 15 12 -3 (1.90) | 15 15 0 (2.96) |
|  | L NAcc | -12 12 0 (2.90)* | -12 6 -3 (2.12) | -15 6 -3 (2.02) |
| Thalamus | R anterior thalamus | 9 -9 3 (4.44) | 9 -9 3 (2.43) | 6 -9 0 (3.32) |
|  | L anterior thalamus | -6 -6 0 (2.50) | n.s. | -3 -6 0 (2.66) |

Note. To reduce false negatives, all regions reaching an uncorrected threshold of $p < .05$ are reported. Asterisks (\*) indicate statistical significance at $p < .05$ , family-wise error (FWE)-corrected for small volumes using a 3-mm radius sphere centered on a priori coordinates for the bilateral nucleus accumbens and ventral tegmental area (Diekhof and Gruber, 2010). Crosses (†) denote a trend toward significance at $p < .10$ (FWE-corrected). R, right; L, left; Mir, mirtazapine treatment group; Ago, agomelatine treatment group; SSRI, selective serotonin reuptake inhibitor group; RS, responders; NRS, non-responders; avPFC, anteroventral prefrontal cortex; IFG, inferior frontal gyrus; MFG, middle frontal gyrus; NAcc, nucleus accumbens; OFC, orbitofrontal cortex; SFS, superior frontal sulcus; TOC, temporo-occipital cortex; VS, ventral striatum; VTA, ventral tegmental area; >, greater activation than; Mir > Ago, (Mir RS > Mir NRS) > (Ago RS > Ago NRS); Mir > SSRI, (Mir RS > Mir NRS) > (SSRI RS > SSRI NRS); n.s., not significant.

Among regions showing prototypical task-related activation, increased activation specifically associated with treatment response to mirtazapine—relative to both the SSRI and agomelatine treatment arms—was primarily observed in prefrontal areas including the right pars opercularis of the inferior frontal gyrus (IFG; Broca’s homologue), the central orbitofrontal cortex (OFC), and the middle third of the middle frontal gyrus (MFG), as well as in the fusiform gyrus. Subcortically, greater activation related to mirtazapine treatment response was observed in the ventral putamen and the VTA (Table 2A).

Further regions showing increased activation related to mirtazapine treatment response, though only partially specific to mirtazapine, included prefrontal areas such as the frontomedian cortex, posterior superior frontal sulcus (SFS), and anteroventral prefrontal cortex (avPFC), in addition to other cortical regions including the intraparietal cortex, temporo-occipital cortex (TOC), and anterior insula (Table 2B). In subcortical areas, increased activation was observed in the nucleus accumbens (NAcc) and the anterior thalamus. These observed differences in neural activation between responders and non-responders were partially specific to mirtazapine, being clearly absent in the SSRI group but either comparable or relatively attenuated in the agomelatine group.

Notably, among the a priori regions, increased activation associated with mirtazapine response in the bilateral NAcc remained significant after small volume correction (*p* < .05, FWE-corrected), while a trend-level effect was noted in the right VTA (*p* = .07, FWE-corrected).

## 4. Discussion

This study aimed to investigate pathophysiological changes in patients with MDD that may be predictive of treatment response to mirtazapine, using pre-treatment fMRI during a conditioned reward task. Compared to non-responders, mirtazapine responders exhibited increased activation across numerous reward-related brain regions, including the VTA and VS, as well as prefrontal, parietal, temporal, and insular cortices. More specifically, increased activation uniquely associated with mirtazapine response—compared to both the SSRI and agomelatine treatment groups—was observed in the VTA, ventral putamen, and prefrontal subregions (Table 2A). This differential activation pattern was completely absent in the other two treatment groups, suggesting clear specificity for mirtazapine treatment response. Further brain regions showed increased activation related to mirtazapine treatment response, which was specific only when compared to the SSRI treatment arm, indicating partial specificity. These included additional prefrontal areas and the NAcc, as well as parietal, temporal, and insular cortices, and the thalamus (Table 2B). While this treatment response-related increase in activation was not seen in the SSRI group, it was either similar or moderately reduced in the agomelatine group. To our knowledge, this is the first study to investigate neurofunctional markers that may be associated with mirtazapine response in MDD using a reward-based task. The findings suggest potential biomarkers for a specific MDD subtype that responds particularly well to mirtazapine and may offer insights into the underlying neurobiological mechanisms of this specific antidepressant treatment response.

### 4.1 Bottom-up dopaminergic pathway alterations

Mirtazapine responders exhibited enhanced activation within key structures of the mesolimbic dopaminergic system, notably the VTA and VS, including the ventral putamen and NAcc. These regions are central to bottom-up dopaminergic signaling and were explicitly designated as a priori regions in our analysis, given the task’s design to engage dopaminergic responsiveness to reward stimuli. They facilitate reward processing through dopaminergic transmission, with inputs originating from the VTA projecting to the VS (Haber and Knutson, 2010).

Consistent with our findings, various forms of dysfunction in these regions have been reported in MDD, particularly during reward-related tasks when compared with healthy controls (Clery-Melin et al., 2019; Goya-Maldonado et al., 2015; Jullig et al., 2024). Specifically, hypoactivation of the VS has been frequently observed across a range of reward-processing tasks (Goya-Maldonado et al., 2015; Pizzagalli et al., 2009; Segarra et al., 2016; Zhao et al., 2024). Although less commonly reported, instances of VS hyperactivation have also been documented (Goya-Maldonado et al., 2015). The VTA, due to its small size and susceptibility to signal distortion, remains less frequently studied using fMRI. Nevertheless, both hypoactivation (Dillon et al., 2014; Goya-Maldonado et al., 2015) and hyperactivation (Kumar et al., 2008) of the VTA during reward processing have been reported in MDD. In parallel, complementary findings from animal models further support the presence of VTA dysfunction in depression (Chaudhury et al., 2013; Tye et al., 2013). Moreover, altered functional connectivity between the VTA and VS has also been observed in MDD, with studies reporting both reduced (Kumar et al., 2018) and increased connectivity (Redlich et al., 2015) during reward-related tasks. Taken together, these findings support the view that dysfunction within the dopaminergic pathway, including the VTA and VS, is one of the core pathophysiological features of MDD (Nestler and Carlezon, 2006; Russo and Nestler, 2013).

Furthermore, the heterogeneity in VTA and VS activation patterns among patients with MDD has been proposed to reflect distinct pathophysiological mechanisms related to the dopaminergic pathway, potentially representing a specific subtype of the disorder. For instance, patients with MDD exhibiting relatively decreased activation in the VTA and VS—or reduced functional connectivity between them—during reward processing have been suggested to experience bottom-up dopaminergic signaling deficits (Goya-Maldonado et al., 2015; Kumar et al., 2018). Conversely, increased connectivity between the VTA and VS during reward processing may reflect compensatory mechanisms, such as postsynaptic dopamine receptor upregulation in response to reduced dopamine release (which is often observed in MDD), or excessive prefrontal regulation that suppresses VS activity (Redlich et al., 2015).

Overall, our finding of increased activation in the VTA and VS among mirtazapine responders may reflect alterations in bottom-up dopaminergic reward processing. However, this pattern may also represent another pathophysiological feature of MDD, as suggested by previous studies potentially indicating relatively preserved bottom-up signaling, an exaggerated response resulting from impaired prefrontal suppression, or compensatory upregulation in response to dopamine insufficiency. Further research is required to clarify these underlying mechanisms. In summary, such enhanced activation during reward processing may characterize a biologically distinct subtype of MDD that is particularly responsive to mirtazapine.

### 4.2 Alterations in higher-order prefrontal regulation of reward processing

Mirtazapine responders exhibited increased activation across several prefrontal regions implicated in the regulation of reward processing, including the OFC, pars opercularis of the IFG, avPFC, MFG (middle third), SFS, and frontomedian cortex. These regions collectively support a range of higher-order regulatory functions that guide reward-related behavior.

The OFC is involved in integrating sensory information and evaluating it in terms of reward value (Rolls, 2019). It transmits this information to other prefrontal regions and modulates reward-related dopaminergic activity in the VTA via top-down reward signaling through basal ganglia structures, including the VS (Haber, 2014; Wallis and Miller, 2003). The pars opercularis of the IFG has been implicated in proactive behavioral inhibition, particularly in response to changing reward contingencies in non-reward contexts (Aron et al., 2014; Deng et al., 2017), by linking the OFC to premotor areas (Rolls et al., 2020). The avPFC contributes to suppressing immediate reward-driven responses in favor of long-term goal pursuit by modulating dopamine-mediated signaling in the VS (Diekhof and Gruber, 2010). Furthermore, the dorsolateral prefrontal regions, including the MFG and SFS, support flexible, context-sensitive reward-guided decision-making by integrating reward value signals from the OFC with spatial attention and task-relevant inputs from other brain areas (Brevers et al., 2023; Wallis, 2007). The frontomedian cortex, associated with self-referential processing, is thought to support self-initiated behavioral control by integrating internal motivational signals (Zysset et al., 2002) and modulating premotor regions to translate these into action (Kuhn et al., 2009; Lynn et al., 2014), rather than relying on externally cued inhibition. Together, these prefrontal regions orchestrate various regulatory functions, such as reward valuation, behavioral inhibition, and goal-directed control, which collectively shape reward-related behavior. The increased prefrontal activation observed among mirtazapine responders may therefore reflect altered regulatory engagement during reward processing.

Supporting this interpretation, a growing body of research has reported abnormal prefrontal activity in MDD across various stages of reward processing. These abnormalities are thought to reflect pathophysiological disruptions in regulatory mechanisms. Notably, the prefrontal regions that showed increased activation in mirtazapine responders in our study substantially overlap with those previously identified as exhibiting aberrant activation during reward processing in MDD (Borsini et al., 2020; Ng et al., 2019; Zhang et al., 2013; Zhao et al., 2024). Importantly, the specific patterns of aberrant activation have varied across studies, even when examining similar regions and stages of reward processing. This variability has been interpreted as indicative of distinct pathophysiological features across MDD subtypes, potentially reflecting heterogeneous disruptions in prefrontal regulatory control (Pizzagalli and Roberts, 2022).

For instance, increased activation of the avPFC and VS during reward processing in patients with MDD has been suggested to reflect dysfunctional top-down inhibition by the avPFC over the VS, which may lead to difficulty suppressing the urge to pursue immediate rewards in favor of long-term goals (Goya-Maldonado et al., 2015; Jullig et al., 2024). On the other hand, a previous meta-analysis (Ng et al., 2019) proposed that OFC hyperactivity may reflect excessive top-down control over subcortical reward systems, potentially leading to a blunted hedonic response. While based on meta-analytic observations, this interpretation is consistent with findings from optogenetic and neuroimaging animal research demonstrating that OFC hyperactivity can suppress dopaminergic signaling in the midbrain and attenuate striatal responses (Ferenczi et al., 2016). Additionally, overactivation of medial and dorsolateral prefrontal regions, accompanied by reduced striatal activation during reward processing, has been interpreted as a compensatory mechanism for diminished striatal responsiveness, which has been associated with anhedonic symptoms in MDD (Pizzagalli and Roberts, 2022).

Thus, our finding of increased prefrontal activation in mirtazapine responders may reflect a distinct pattern of altered regulatory processing in MDD. Given that this activation spanned multiple prefrontal regions involved in diverse aspects of reward-related regulation, including reward valuation, behavioral inhibition, and goal-directed control, it likely represents functional alterations across several regulatory processes implicated in MDD pathophysiology. Collectively, these findings raise the possibility that this neural activation profile during reward processing may serve as a neural signature of an MDD subtype more likely to benefit from mirtazapine.

### 4.3 Alterations in supportive regions of prefrontal reward regulation

In addition to prefrontal regions, mirtazapine responders exhibited increased activation in several brain areas functionally connected to the prefrontal cortex, including the anterior and posterior insula, fusiform gyrus, TOC, anterior thalamus, and intraparietal cortex. These regions are believed to support reward-related behavior by relaying various types of information essential for higher-order prefrontal regulation.

The insular cortex receives bodily-state inputs—including gustatory, somatosensory, and interoceptive signals—from peripheral sensory pathways, which are initially processed in the posterior insula. This information is then integrated in the anterior insula with affective and cognitive signals from limbic and prefrontal regions to generate subjective feeling states related to reward. These signals support the OFC in reward evaluation and aid the dorsolateral prefrontal cortex in regulating reward-guided behavior by conveying bodily-state and affective information (Levy and Glimcher, 2012; Namkung et al., 2017). The fusiform gyrus and TOC, both parts of the ventral visual stream, are involved in category-specific recognition (e.g., of faces and objects) (Ishai et al., 1999; Kanwisher et al., 1997) and in integrating semantic and affective features of visual stimuli (Abdel-Ghaffar et al., 2024), respectively. These regions support the OFC’s role in reward evaluation by providing processed visual inputs. The intraparietal cortex, a component of the dorsal visual stream, encodes spatial information and prioritizes attention accordingly (Corbetta and Shulman, 2002; Vossel et al., 2014), while the anterior thalamus contributes to adaptive decision-making by maintaining and updating contextual and rule-based information through interactions with limbic and cortical structures (Nelson, 2021). Together, these two regions support flexible, reward-based decision-making in the dorsolateral prefrontal cortex by supplying spatial, attentional, and task-relevant inputs that are integrated with reward value signals from the OFC.

In line with this, abnormal functioning of these regions has previously been implicated in MDD and is thought to reflect underlying pathophysiological features of the disorder. During reward processing, hypoactivation of the insula (Pizzagalli et al., 2009; Zhang et al., 2013) and thalamus (Reinen et al., 2021; Segarra et al., 2016; Zhang et al., 2013) has been reported in individuals with MDD compared with healthy controls. Conversely, hyperactivation has been observed in the fusiform gyrus (Pizzagalli et al., 2009; Smoski et al., 2009; Zhang et al., 2013; Zhao et al., 2024), TOC (Smoski et al., 2009), and intraparietal cortex (Zhang et al., 2013).

Altogether, our finding of increased activation in the anterior and posterior insula, anterior thalamus, fusiform gyrus, TOC, and intraparietal cortex—regions that support prefrontal regulation by processing and relaying various types of reward-related information, including affective, interoceptive, visual, contextual, and attentional signals—may reflect the functional engagement of distributed prefrontal regulatory networks, extending beyond the prefrontal cortex, during reward processing in mirtazapine responders. This activation pattern may indicate altered functioning within broader prefrontal-associated regulatory systems, potentially relevant to the pathophysiology of MDD. As such, this neurofunctional profile may serve as a potential marker of enhanced responsiveness to mirtazapine.

### 4.4 Specificity of reward-related activation patterns to mirtazapine treatment response

Interestingly, some of the increased activation patterns associated with treatment response in the mirtazapine group—observed in regions such as the frontomedian cortex, avPFC, intraparietal cortex, TOC, anterior and posterior insula, anterior thalamus, and NAcc—were only partially specific to mirtazapine. Although these activation patterns were absent in the SSRI group, similar or attenuated activation differences associated with treatment responses were found in the agomelatine group. This observation is consistent with previous findings suggesting that mirtazapine shares greater pharmacological and clinical similarities with agomelatine than with SSRIs.

Unlike the SSRIs used in our study (sertraline and escitalopram), both mirtazapine and agomelatine antagonize 5-HT_2C_ receptors. This mechanism is believed to enhance noradrenergic and dopaminergic transmission, which play crucial roles in reward-related processing (Millan et al., 2003; Zhang et al., 2023). Additionally, each antidepressant is known to alleviate sleep disturbances in patients with MDD, although via distinct mechanisms: mirtazapine through histaminergic H1 receptor antagonism (Davis, 2020) and agomelatine through melatonergic MT_1_/MT_2_ receptor agonism (Manikandan, 2010). These shared pharmacological and clinical properties, which differ from those of SSRIs, may help explain the partial overlap of increased brain activations associated with treatment responses to both mirtazapine and agomelatine—but not to SSRIs. Moreover, the clearly distinct activation profile associated with treatment response in the mirtazapine group, compared with the SSRI group, likely reflects mirtazapine’s unique pharmacological actions, including its combined antagonism of 5-HT_2_/5-HT_3_, α_2_-adrenergic, and H_1_ receptors, in contrast to the serotonergic reuptake inhibition mechanism of SSRIs. In this context, our findings also suggest that patients with MDD who respond to agomelatine share, at least in part, similar pathophysiological characteristics with those who respond to mirtazapine, particularly involving disturbances in bottom-up processing and regulatory control within the reward system.

### 4.5 Previous fMRI research on mirtazapine response

To date, no studies have directly examined the association between pre-treatment neural activation during reward processing and treatment response to mirtazapine in patients with MDD, as investigated in the present study. fMRI research on mirtazapine has been limited, with only one prior study employing a reward-related task, which was conducted in healthy individuals. It reported increased activation in the parietal cortex during reward processing following mirtazapine administration (Vollm et al., 2006). Conversely, fMRI studies involving patients with MDD have primarily employed emotion-related tasks, such as emotional face-matching paradigms. For example, one study found that reduced activation in the rolandic operculum during implicit emotion processing was associated with a better treatment response to mirtazapine (Frodl et al., 2011). In a related study using the same paradigm, increased functional connectivity between the OFC and dorsolateral prefrontal cortex during emotional face processing was reported following mirtazapine treatment (Lisiecka et al., 2011). Although the specific regions identified in these studies do not entirely overlap with those differentiating responders from non-responders in our analysis, several key areas, including the parietal cortex, OFC, and dorsolateral prefrontal cortex, are common to both prior findings and the present study. These overlaps, particularly within the prefrontal regulatory network, suggest a degree of consistency across studies and provide tentative support for the neural activation patterns associated with the mirtazapine response observed here.

While these parallels support the plausibility of our findings, several factors may account for the discrepancies between our results and previous studies, including differences in task paradigms (e.g., reward-based decision-making tasks versus emotional recognition tasks), the timing of neural measurement (e.g., pre-treatment versus post-treatment), and study populations (e.g., healthy individuals versus patients with MDD).

Since brain activation patterns are shaped by and responsive to the specific task paradigm, differences in task design can naturally lead to variations in observed neural activity. Notably, post-treatment fMRI studies reflect medication-induced changes in brain function, which may differ from pre-treatment activation patterns, as assessed in our study. In addition, because healthy individuals are less likely to experience brain dysfunctions observed in MDD, their activation profiles may differ substantially from those of patients. Further research employing diverse fMRI paradigms and incorporating direct comparisons between treatment responders and healthy controls is needed to clarify the underpinnings of mirtazapine responsiveness. Such efforts will deepen our understanding of an MDD subtype responsive to mirtazapine and support the development of robust, integrated biomarkers for predicting treatment outcomes.

### 4.6 Limitations

Our study has some limitations that should be addressed in future research. First, most treatment groups included a relatively small number of participants, which may have limited the statistical power and generalizability of the findings. Despite this, the results are robust and consistent with a substantial body of prior research highlighting the central role of the reward system in the pathophysiology of MDD (Jiang et al., 2023; Ng et al., 2019; Oh et al., 2023; Treadway and Pizzagalli, 2014). Moreover, we verified that the identified response-related activation patterns overlapped with prototypical task-related activation, as defined by a reference map derived from a large healthy control sample from the GIG cohort. This overlap indicates that the activations occurred within task-relevant regions, reducing the likelihood that they reflect random or non-specific background signals and thus supporting the interpretability of the results despite the modest sample size. Second, since we did not include a healthy control group for comparison, the activation patterns observed in mirtazapine responders vs. non-responders cannot be unambiguously attributed to either MDD-related abnormalities or typical neural responses to the reward task. Nevertheless, the distinctiveness of these patterns relative to other treatment groups reinforces their interpretation as potential neurofunctional features unique to the mirtazapine-responsive subtype of MDD. Future studies with larger samples and the inclusion of healthy control comparisons may enhance the interpretability and generalizability of these findings.

## 5. Conclusion

In conclusion, this study is the first to investigate pathophysiological changes of the extended reward system in patients with MDD that may be predictive of the treatment response to mirtazapine. The distinct brain activation patterns observed in the mirtazapine responders may serve as potential neuropathophysiological biomarkers for an MDD subtype that is particularly responsive to mirtazapine. Drawing on extensive neuroimaging research on MDD and reward processing, our findings suggest that mirtazapine responders may exhibit alterations not only in the bottom-up dopaminergic pathway but also particularly in multiple prefrontal control networks involved in reward evaluation, behavioral inhibition, and reward-based decision-making. By shedding light on the neural mechanisms underlying the mirtazapine response, our study contributes to the advancement of biomarker-based approaches in antidepressant selection, thereby aiding the progression of precision medicine in MDD. Future studies incorporating larger samples, healthy control comparisons, and a wider range of task paradigms, including both reward-based and emotion-related processes, are warranted to identify clinically meaningful neural signatures that inform treatment selection and to establish reliable biomarkers for treatment prediction in MDD.

## Data Availability

Processed data supporting the findings of this study are available from the corresponding author upon reasonable request.

## Acknowledgment

We thank Helena Metzker, Bernd Krämer, Miriam Pfister, Lennart Berzow, and Lea Winter for their support in data handling and processing.

