## Supplementary Table 1 for "Neuropathophysiological changes associated with mirtazapine treatment response in major depressive disorder: insights into bottom-up dopaminergic pathways and prefrontal control networks"

| Supplementary table 1. Decreased deactivation in responders to mirtazapine relative to non-responders | | |
| --- | --- | --- |
|  | Region | Mir RS > NRS |
|  |  | MNI coordinate (T-value) |
| Frontal | L pregenual ACC | −12 57 6 (2.82) |
|  | R anterior inferior frontal sulcus | 42 36 3 (2.66) |
|  | R posterior middle frontal gyrus | 42 24 42 (2.61) |
|  | L subgenual ACC | −9 33 −3 (2.44) |
|  | R pregenual ACC | 6 48 9 (2.15) |
| Parietal | R posterior inferior parietal cortex | 33 −66 48 (2.73) |
| Temporal | R parahippocampal gyrus | 15 −33 −15 (2.96) |
|  | L parahippocampal gyrus | −27 −33 −9 (2.40) |
|  | R middle temporal gyrus | 60 −45 9 (2.86) |
|  | L middle temporal gyrus | −66 −39 0 (2.72) |
|  | R posterior superior temporal gyrus | 45 −33 9 (2.62) |
|  | L superior temporal gyrus | −48 −12 6 (2.59) |
| Occipital | R O4 | 36 −69 −6 (3.84) |
|  | R visual cortex | 33 −6 9 (3.72) |
| Subcortical | L pulvinar thalami | −15 −27 3 (3.20) |
|  | R medial thalamus | 3 −21 0 (3.09) |
|  | L cerebellum | −15 −33 −21 (2.57) |
|  | R posterior putamen | 18 −45 6 (2.56) |
|  | L posterior ventral putamen | −24 −6 −9 (2.22) |
| *Note. To reduce false negative results, we included all brain regions reaching a minimum *p* < .05 (uncorrected). R, right; L, left; ACC, anterior cingulate cortex; **Mir**, mirtazapine treatment group; **RS**, responders; **NRS**, non-responders; **>**, greater activation than. | | |
